# A *de novo* missense variant in *EZH1* associated with developmental delay exhibits functional deficits in *Drosophila melanogaster*

**DOI:** 10.1101/2023.01.31.23285113

**Authors:** Sharayu Jangam, Lauren C. Briere, Kristy Jay, Jonathan C Andrews, Melissa A. Walker, Lance H. Rodan, Frances A. High, Undiagnosed Diseases Network, Shinya Yamamoto, David A. Sweetser, Michael Wangler

## Abstract

*EZH1* (*Enhancer of Zeste, homolog 1)*, a Polycomb Repressive Complex-2 (PRC2) component, is involved in a myriad of cellular processes through modifying histone 3 lysine27 (H3K27) residues. *EZH1* represses transcription of downstream target genes through H3K27 trimethylation (H3K27me3). Genetic mutations in histone modifiers have been associated with developmental disorders, while *EZH1* has not yet been linked to any human disease. However, the paralog *EZH2* is associated with Weaver syndrome. Here we report a previously undiagnosed individual with a novel neurodevelopmental phenotype identified to have a *de novo* variant in *EZH1*, p.Ala678Gly, through exome sequencing. The individual presented in infancy with neurodevelopmental delay and hypotonia and was later noted to have proximal muscle weakness. The variant, p.A678G, is in the SET domain, known for its methyltransferase activity, and was the best candidate variant found in the exome. Human *EZH1*/*2* are homologous to fly *Enhancer of zeste E(z)*, an essential gene in flies, and the residue (A678 in humans, A691 in *Drosophila*) is conserved. To further study this variant, we obtained *Drosophila* null alleles and generated transgenic flies expressing wild-type *(E(z)*^*WT*^) and the variant *(E(z)*^*A691G*^). The *E(z)*^*A691G*^ variant led to hyper H3K27me3 while the *E(z)*^*WT*^ did not, suggesting this is as a gain-of-function allele. When expressed under the tubulin promotor *in vivo* the variant rescued null-lethality similar to wild-type but the *E(z)*^*A691G*^ flies exhibit bang sensitivity and shortened lifespan. In conclusion, here we present a novel *EZH1 de novo* variant associated with a neurodevelopmental disorder. Furthermore, we found that this variant has a functional impact in *Drosophila*. Biochemically this allele leads to increased H3K27me3 suggesting gain-of-function, but when expressed in adult flies the *E(z)*^*A691G*^ has some characteristics of partial loss-of-function which may suggest it is a more complex allele *in vivo*.

## Introduction

*EZH1* (*Enhancer of Zeste, homolog 1)*, encodes an enzyme with *Histone-lysine N-methyltransferase activity*, which is a component of the histone modifier family Polycomb Repressive complex 2 (PRC2) (Shen et al. 2008; Jacobs and van Lohuizen 1999). The PRC2 complex plays a crucial role in establishing and maintaining chromatin structure by binding to histone tails and adding methyl or acetyl groups, thus tightening or loosening chromatin (Margueron and Reinberg 2011; Kuzmichev et al. 2002; Jacobs and van Lohuizen 1999). Tri-methylation of the histone core’s N-terminus tail often leads to the reorganization of chromatin and the resultant repression of downstream genes (Bannister et al. 2002; Rea et al. 2000). The PRC2 complex comprises proteins including methyl transferases like EZH1/ Enhancer of Zeste, homolog 2 (EZH2), acetyltransferases like Embryonic Ectoderm Development (EED), DNA binding proteins like Suppressor of Zeste 12 (SUZ12), and histone binding proteins like retinoblastoma binding proteins 46/48 (RbAp46/48) (Margueron and Reinberg 2011). In human, *EZH1* and its paralog *EZH2* share 63% identity with each other (Yu et al. 2019) and alter each other in the PRC2 complex (Kook et al. 2017).

EZH1 is involved in the trimethylation of lysine (K) 27 of the Histone 3 (H3) (Shen et al. 2008; Hidalgo and Gonzalez 2013). Tri-methyl groups (me3) are added to H3K27, requiring the SET domains of EZH1. The SET domain consists of 130-140 amino acids and was first characterized in the *Drosophila melanogaster* proteins Su(var)3-9, Enhancer of Zeste E(z), and Trithorax domain, thus named the “SET” domain (Jones et al. 1998; Rastelli et al. 1993). Indeed, lysine methyltransferases have been among the prominent genes identified by classic *Drosophila* studies on position-effect variegation in which changes in the euchromatin or heterochromatin border lead to suppression or enhancement of white gene variegation in the fly eye (Elgin and Reuter 2013; Dillon et al. 2005) (Herz et al. 2013). *E(z)* fly mutants display early embryonic lethality with distinct chromatin abnormalities (O’Dor et al. 2006). Mutant clones of *E(z)*, in the third instar larval brain, have been shown to lead to reduced neuroblast size and reduction in mitotic activity (Bello et al. 2007). In the adult brain, mutant clones lead to misguided axons and/or ectopic glia-like cells (Wang et al. 2006).

Human Mendelian disorders have been reported for multiple members of the PRC2 complex (Deevy and Bracken 2019). Cohen-Gibson syndrome is an overgrowth syndrome with neurodevelopmental abnormalities and dysmorphic features caused by *de novo* missense variants in the *EED* gene (Cohen and Gibson 2016). These variants disrupt EED-EZH1/2 protein interactions (Imagawa et al. 2017). Imagawa-Matsumoto syndrome is an overgrowth syndrome due to variants in SUZ12 (Imagawa et al. 2017), and *EZH2* is responsible for Weaver syndrome, an overgrowth and developmental condition (Weaver et al. 1974).

While *EZH2* and other members of the complex have been linked to overgrowth and developmental delay, *EZH1* is not yet associated with human disease. *EZH1* and *EZH2* are approximately 63% identical and share 94% identity within the SET domain (Yu et al. 2019). Still, their functions are very divergent (Shen et al. 2008). Knock-out of *Ezh2* is lethal in mice (O’Carroll et al. 2001), while heterozygotes have increased growth and body weight, an interesting correlation to the human Weaver overgrowth syndrome (Béguelin et al. 2013). *Ezh1* in mice is not essential and does not associate with obvious overgrowth, but it is essential for hematopoietic stem cell maintenance (Hidalgo et al. 2012; Shen et al. 2008).

Here we report an individual with a *de novo* variant in *EZH1* and a neurodevelopmental phenotype with variant functional studies completed using the *Drosophila* model organism.

## Results

### 1. Case Presentation

{the entire case history is removed as per medRxiv rules. Please contact authors to access the details} Clinical quad exome sequencing, including the patient’s unaffected older sibling, did not reveal pathogenic or likely pathogenic variants that would explain the phenotype; however, subsequent research-based analysis of the data highlighted a novel *de novo* missense variant (NM_001991.3:c.2033G>C (p.A678G)) in *EZH1*, a gene with no established disease association. This finding was Sanger confirmed in a CLIA lab.

### 2. *EZH1* candidate disease-causing variant

The *de novo* missense variant in *EZH1* is considered as a strong novel disease candidate for this case for multiple reasons. *EZH1* is highly expressed in neuronal cells (Uhlén et al. 2015). The variant has not been seen in gnomAD previously (Karczewski et al. 2020). The gene does not have a known relationship to human disease, and has a pLI of 0.04, indicating no constraint for loss of function variants, but the missense Z-score of 4.2 indicates a missense constraint in gnomAD (Karczewski et al. 2020).

Computational predictions suggest the missense variant is deleterious (SIFT = 0.0) (Ng and Henikoff 2001) and probably damaging (Polyphen = 1) (Adzhubei et al. 2010), with a CADD score of 32 (Rentzsch et al. 2021). It falls within the SET domain of EZH1, and the region and residue are highly conserved across species (**FIGURE 2A, B**). EZH1 has been reported to be a repressor of hematopoietic multipotency in the early mammalian embryo (Vo et al. 2018). EZH1 is part of the PRC2 complex, and EZH2, the human paralog and alternative subunit, is known to cause autosomal dominant Weaver syndrome (Imagawa et al. 2017; Weaver 2019). Indeed, the equivalent variant (p.A677G in *EZH2*, aligned with p.A678G in *EZH1*, **FIGURE 2B’**) is a known disease-causing variant in Weaver syndrome (p.A677G {isoform EZH2-Q15910-1} or p.A682G {isoform EZH2-Q15910-2}) (Diets et al. 2018; UniProt Consortium 2022). Moreover, another variant impacting the same amino acid has also been identified as disease-causing in Weaver syndrome (*EZH2* p.A677T or *EZH2* p.A682T) (Tatton-Brown et al. 2013, 2011). Taken together the *EZH1* variant is a strong candidate variant for this case and was therefore submitted to the Model Organisms Screening Center (MOSC) of the UDN and tested in *Drosophila* (Baldridge et al. 2021)

**Figure1:**
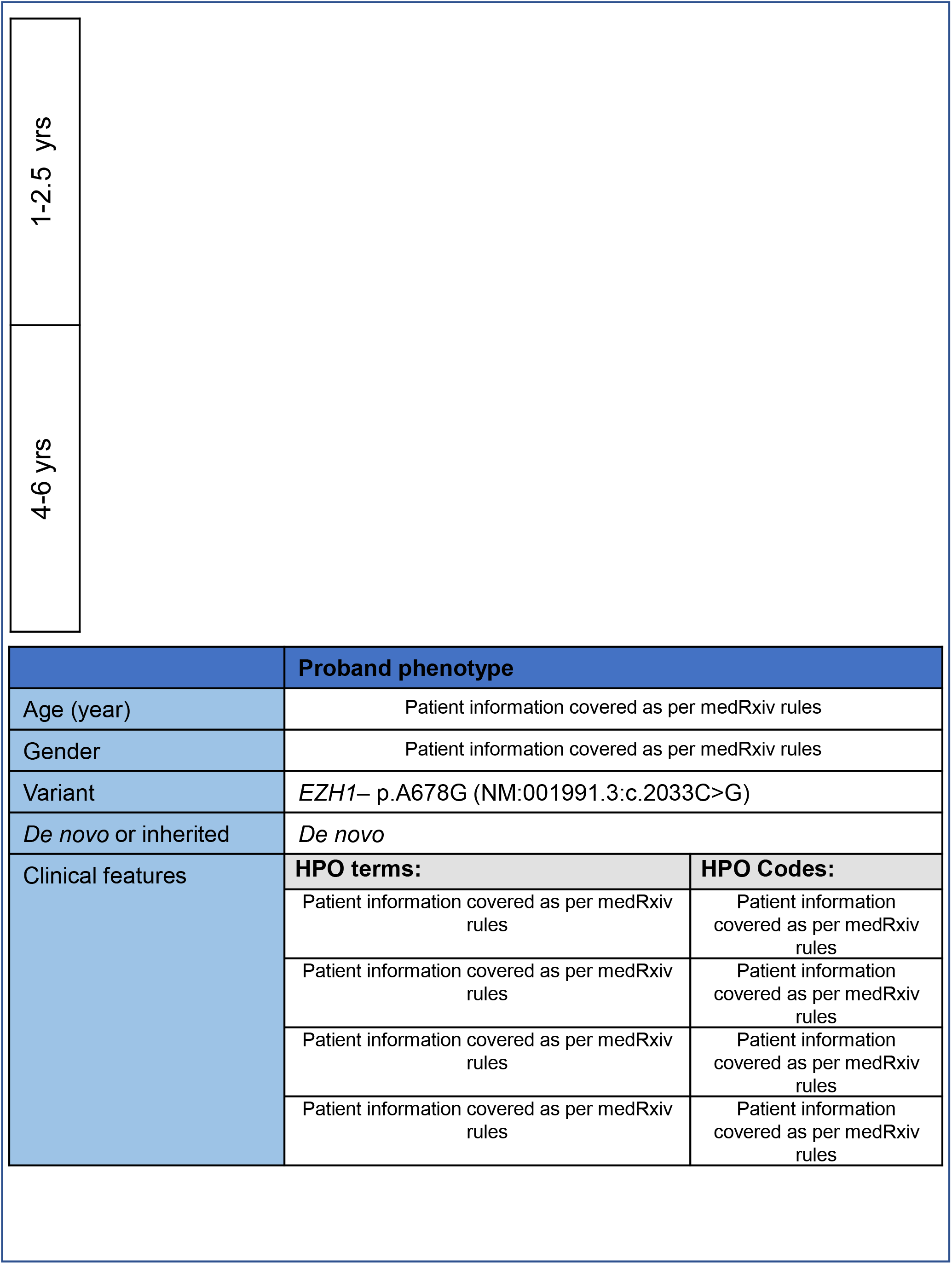
Patient information: **Photos** and clinical features: Patient information is covered as per medRxiv rules please contact authors for the further information.

**Figure 2:**
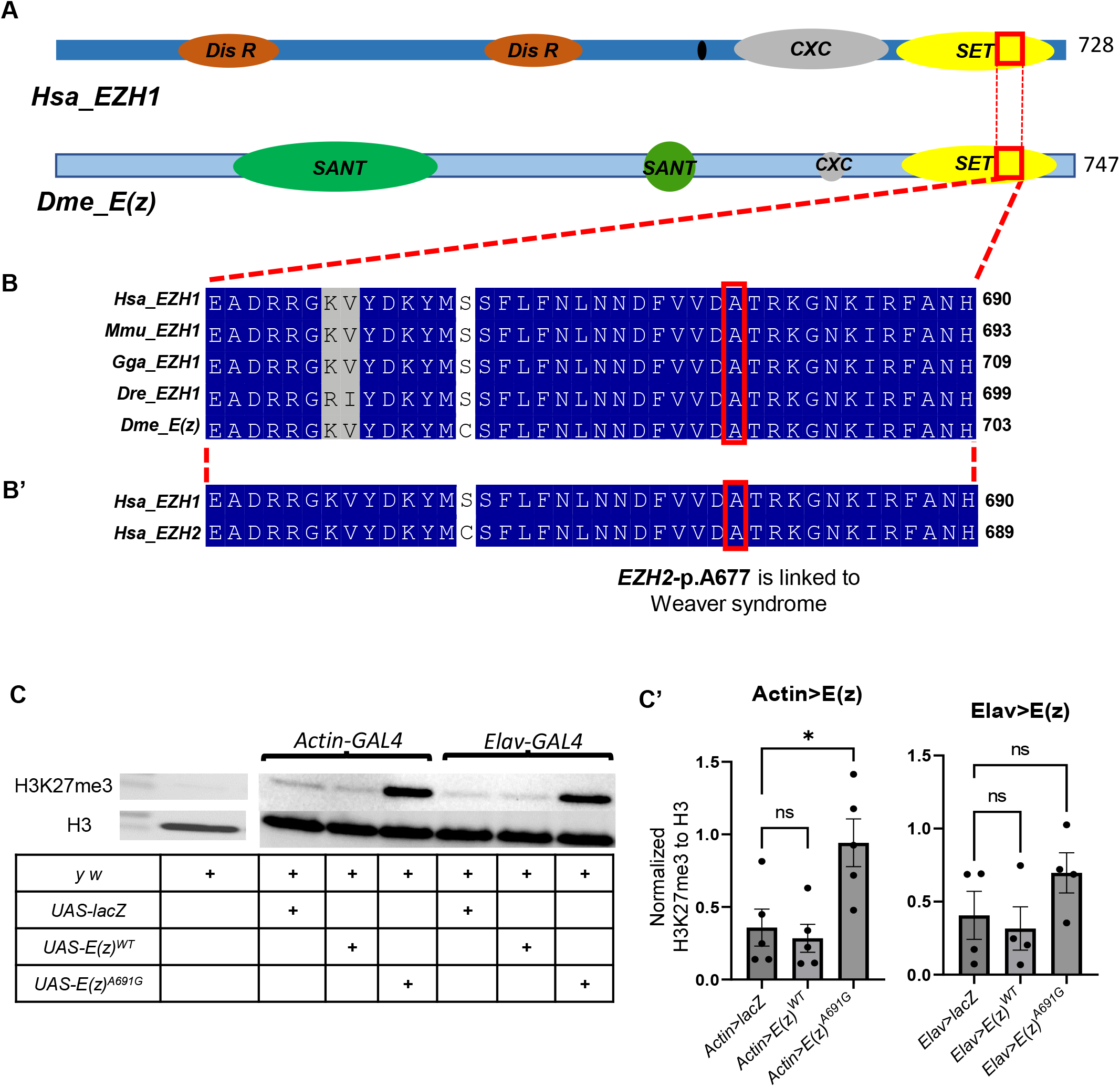
**A: Domain structure schematic of human *EZH1*(*Hsa_EZH1*)** and ***Drosophila melanogaster E(z)(Dmel_E(z))*** **B-B’: The conservation of *EZH1-pA678* throughout the species** and also in both **human paralogs *EZH1* and *EZH2*** (The structures are derived from UniProt.) [*Hsa_EZH1*: Homo Sapiens *EZH1*; Dis R: Disordered Region; SANT: SANT-Myb-Protein binding Domain, CXC: cysteine rich DNA binding Domain; *Dme_E(z)* : *Drosophila melanogaster Enhancer of Zaste (E(z)*] **C-C’: Western blot assay for H3K27me3 of UAS-*E(z)***^***wt***^ **and UAS-*E(z)***^***A691G***^ **overexpression**. Tri-methylation of H3K27 was evaluated in the GAL4 driven *E(z)*^*WT*^ and *E(z)*^*A691G*^ progenies. **C’** is the graphical representation of the normalised H3K27 values to H3 values. *UAS-LacZ* was used as a positive control. p-values were determined by unpaired t-tests.

### 3. EZH1/EZH2 are human orthologs of Drosophila Enhancer of Zeste E(z)

The *EZH1* variant was accepted for modeling in the MOSC Fly Core, and previous model organism phenotypes were reviewed (Wang et al. 2019b, 2017, 2019a). We also obtained resources from prior *Drosophila E(z)* studies. Human *EZH1* and *EZH2* have a single orthologue *E(z)* in *Drosophila melanogaster. E(z)* has greater sequence similarity to *EZH2* than to *EZH1*, (DIOPT 14/15, and 11/16, respectively) (Hu et al. 2011). We obtained *E(z)*^*731*^ (Müller et al. 2002) and *E(z)*^*63*^ (Carrington and Jones 1996), both null alleles. Hetero-allelic knock-out of *E(z)* in flies has been shown to be lethal (Steffen et al. 2013), and we confirmed *E(z)*^*731*^*/E(z)*^*63*^ is lethal **(FIGURE 2A)**. The *de novo* p.A678G variant in *EZH1* is found within a highly conserved region within a stretch of the SET domain. In *Drosophila* it is *E(z)* p.A691G **(FIGURE 2B)**.

### 4. A678G variant transgenics in *Drosophila*

We generated the *Drosophila E(z)* cDNA wild-type and variant transgenic flies under the UAS promoter, the *UAS-E(z)*^*WT*^ and *UAS-E(z)*^*A691G*^ flies (**Supplementary FIGURE1A**). Overexpression of these constructs with different *GAL4* lines did not show any noticeable phenotypic differences between WT and variant (**Supplementary Table FIGURE1B**). We evaluated H3K27 trimethylation (H3K27me3) in a western blot assay and noted significantly increased H3K27me3 in *Actin-GAL4>UAS-E(Z)*^*A691G*^ flies (**FIGURE2C-C’**).

We also generated UAS transgenics for human *EZH1* reference and A678G variant flies (**Supplementary FIGURE2**). We expressed the human proteins ubiquitously using a ubiquitous *Actin-GAL4* driver and a neuronal-specific *Elav-GAL4* driver. We did not observe any phenotypic defects as the product of the expression of variant or reference lines. When the overexpressed progenies were tested biochemically for H3K27me3, no significant difference was observed in the level of trimethylation. Moreover, these flies fail to rescue the lethality from an *E(z)-RNAi* knockdown (BL#36068 y^1^ sc* v^1^ sev^21^; p{y^+t7.7^ v^+t1.8^=TRiP.GL00486}attP40) with *Actin-GAL4*. We, therefore, studied the conserved variant in the fly *E(z)*-gene.

### 5. *E(z)*^*A691G*^ rescues the heteroallelic lethality

Previous work generated a constitutively active EGFP tagged *E(z)-cDNA* construct under the control of the tubulin promoter [ptub::*EGFP::E(z)]*, which rescued the heteroallelic lethality of *E(z)* (Steffen et al. 2013). We obtained this construct and used site-directed mutagenesis to generate the p.A691G variant (analogous to *EZH1* p.A678G) and generated transgenic animals of wild-type and variant constructs (**Supplementary FIGURE S3)**. We used these constructs to rescue null alleles previously obtained (*E(z)*^*731*^ (Müller et al. 2002) and *E(z)*^*[63]*^ (Carrington and Jones 1996)). These alleles cause homozygous lethality and fail to complement each other thus resulting in heteroallelic lethality (**FIGURE 3A**). ptub::*E(z)*^*WT*^ and variant ptub::*E(z)*^*A691G*^ were expressed in a heteroallelic background (*E(z)*^*731*^/*E(z)*^*63*^). Introduction of these transgenic lines containing ptub::*E(z)*^*WT*^ or ptub::*E(z)*^A691G^ fully rescued the lethality (**FIGURE 3A**). Moreover, the Mendelian ratios were similar when both constructs were expressed (**FIGURE 3A’**).

**Figure3:**
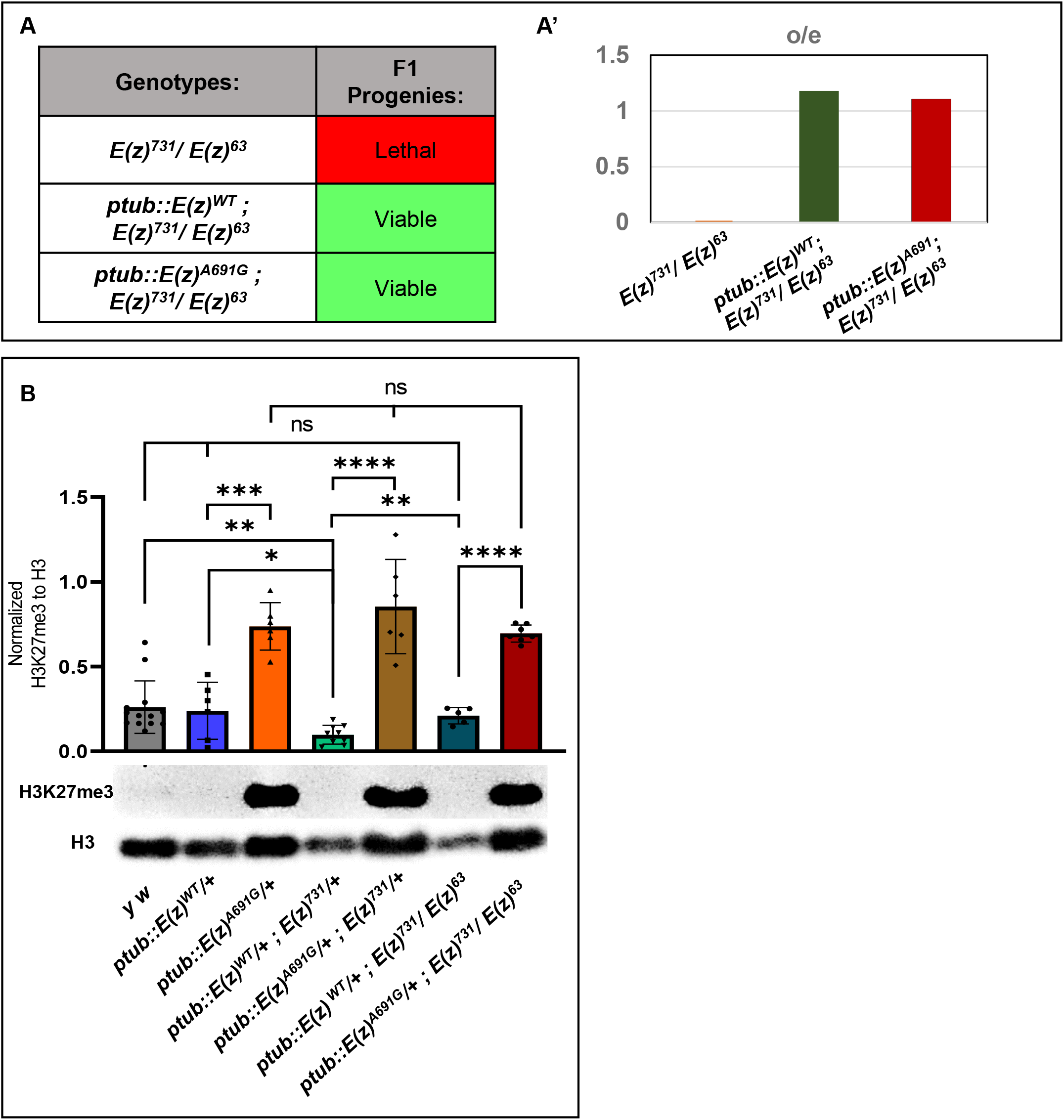
Functional assessment of the transgenic flies: **A-A’. Genetic crosses for the rescue of heteroallelic lethality:** Under constitutively active tubulin promotor (ptub), the WT and the Variant constructs, were both rescued the *E(z)*^*731*^*/ E(z)*^*63*^ heteroallelic lethality (Table A). A’ represents the graphical representation of the observed/expected ratio of the F1 progenies explained in Table A. **B: Western blot assay for H3K27-trimethylation**: Under constitutively active tubulin promotor, the H3K27me3 levels in the variant were compared to WT (p=0.0002). In the sensitized background, when one copy of *E(z)* is removed using a null allele (*E(z)*^*731*^), the variant continues to show hyper tri-methylation as compared to WT (p<0.0001). Even in the fly null background, when compared with WT, the variant maintains the hyper-trimethylation (p<0.0001). p-value were determined by unpaired t-tests.

### 6. *E(z)*^*A691G*^ causes hyper trimethylation of H3K27

*E(z)*, a methyltransferase, has been shown to affect H3K27 methylation (Stepanik and Harte 2012; Cao et al. 2002). We, therefore, tested whether adult flies also show differences in H3K27me3. Introduction of ptub::*E(z)*^*WT*^ [*ptub::E(z)*^*WT*^*/+*] does not impact H3K27me3, but introduction of the variant ptub::*E(z)*^*A691*^ [*ptub::E(z)*^*A691G*^*/+*] leads to ∼ 2-fold increase in H3K27me3 (**FIGURE 3B**, p=0.0002). In a heterozygous null background, this difference was more pronounced. When the wild-type transgenic construct in a heterozygous null background was evaluated [*ptub::E(z)*^*WT*^*/+*; *E(z)*^*731*^*/+*] with control fly strain [*y w*], it appeared to have slightly reduced trimethylation [p=0.073]. But the variant *ptub::E(z)*^*A691G*^ in the same background continues to cause hyper-trimethylation [*ptub::E(z)*^*A691G*^*/+*; *E(z)*^*731*^*/+*] compared to wild-type (**FIGURE 3B**, p<0.0001). Additionally, as both the transgenes rescue the *E(z)*^*731*^*/E(z)*^*63*^ null lethality, we quantified the difference in H3K27me3 levels in these flies. The wild-type [*ptub::E(z)*^*WT*^*/+*; *E(z)*^*731*^*/E(z)*^*63*^] has normal trimethylation levels compared to *y w*, but the variant [*ptub::E(z)*^*A691G*^*/+*; *E(z)*^*731*^*/E(z)*^*63*^] continues to show abnormally high trimethylation compared to wild-type (**FIGURE 3B**, p<0.0001). The variant transgene ptub::*E(z)*^A691G^ maintains hyper-trimethylation in the different genetic backgrounds and suggests a gain-of-function variant in this biochemical assay

### 6. *E(z)*^*A691G*^*-*rescued flies display early bang sensitivity and reduced longevity

The patient has severe global developmental delay, suggesting neurological and/ or neurodevelopmental deficits. Given that the transheterozygote lethality of *E(z)*^*731*^*/E(z)*^*63*^ can be rescued by both ptub::*E(z)*^*WT*^ and the variant ptub::*E(z)*^*A691G*^ and given that ptub::*E(z)*^*A691G*^ has increased H3K27me3, we assessed if ptub::*E(z)*^*A691G*^ causes any phenotype in adult flies. It has been shown that clones expressing the null allele *E(z)*^*731*^ have misguided axons and/or ectopic glia-like cells in adult flies (Wang et al. 2006). To test any potential neurological phenotypes, we performed a bang sensitivity assay at 5 days after eclosion (DAE) and 16 DAE (+/-1 days). When tested in the fly null background at 5 DAE, the variant ptub::*E(z)*^*A691G*^ [*ptub::E(z)*^*A691G*^*/+*; *E(z)*^*731*^*/E(z)*^*63*^] showed significant bang-sensitivity when compared to ptub::*E(z)*^*WT*^ [*ptub::E(z)*^*WT*^*/+*; *E(z)*^*731*^*/E(z)*^*63*^] or *y w* (**FIGURE 4A**, p=0.0041). To test if these transgenes have any effect on bang sensitivity by themselves, the same assay was performed in the fly wild-type background. At 5DAE, neither the ptub::*E(z)*^*A691G*^ [*ptub::E(z)*^*A691G*^*/+*] nor the ptub::*E(z)*^*WT*^ [*ptub::E(z)*^*WT*^*/+*] flies exhibited bang sensitivity compared to *y w* (**FIGURE 4A’**, p=ns). However, when the bang sensitivity assay was performed at 16 DAE in the variant in the fly null background ptub::*E(z)*^*A691G*^ [*ptub::E(z)*^*A691G*^*/+*; *E(z)*^*731*^*/E(z)*^*63*^] again showed significant bang sensitivity compared to wild-type [*ptub::E(z)*^*WT*^*/+*; *E(z)*^*731*^*/E(z)*^*63*^] (**FIGURE 4B**, p=0.0004). Interestingly, at 16 DAE, transgenic ptub::*E(z)*^*A691G*^ flies in a wild-type background [*ptub::E(z)*^*A691G*^*/+*] were seen to be bang sensitive when compared to ptub::*E(z)*^*WT*^ [*ptub::E(z)*^*WT*^*/+*] (**FIGURE 4B’**, p=0.0002).

**Figure4:**
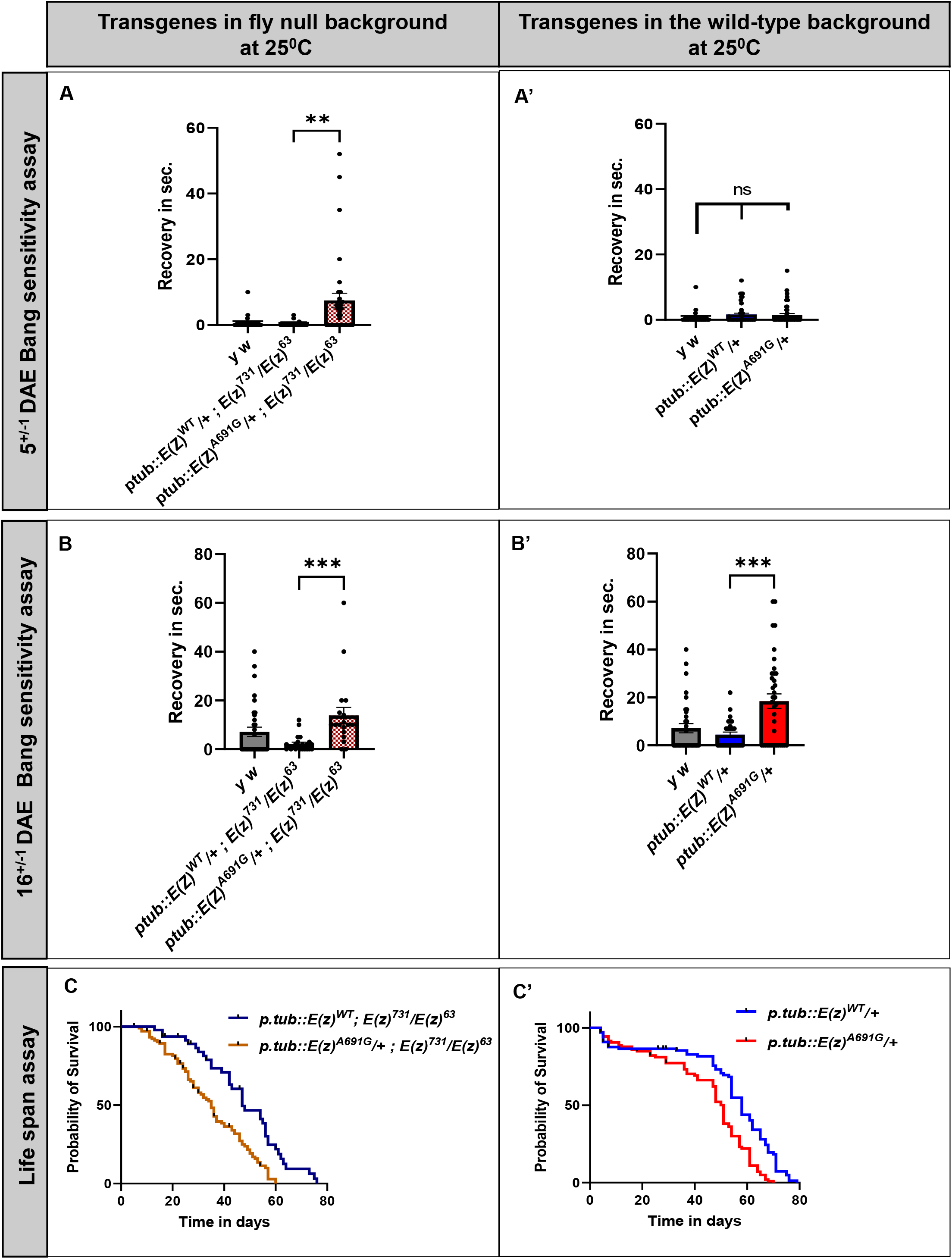
Behavior study with the transgenic flies. **A-A’: Bang sensitivity performed at 5 days after eclosion (DAE):** ptub::*E(z)*^*A691G*^ transgenic flies in the fly null background observed to be bang sensitive than ptub::*E(z)*^*WT*^ (**FIGURE4A**, p=0.0041). When similar experiment is carried out in the wild-type background ptub::*E(z)*^*A691G*^ do not behave different than the wild-type ptub::*E(z)*^*WT*^ (**FIGURE4A’**, p=ns). **B-B’: Bang sensitivity performed at 16 DAE:** ptub::*E(z)*^*A691G*^ transgenic flies in the fly null background continues to show bang sensitivity at 16 DAE when compared to ptub::*E(z)*^*WT*^ (**FIGURE4B**, p=0.0004). In the wild-type background the *ptub::E(z)*^*A691G*^ starts showing the bang sensitivity when compared to ptub::*E(z)*^*WT*^ (**FIGURE4B’**, p=0.0002). **C-C’: Life span assay** : ptub::*E(z)*^*A691G*^ transgenic flies in the fly null background live shorter than the ptub::*E(z)*^*WT*^ (**FIGURE4C, Median Survival: *E(z)***^***WT***^ **= 47, *E(z)***^***A691G***^ ***=* 35**, p<0.0001). However, in the fly wild-type background, ptub::*E(z)*^*A691G*^ transgenic flies shows a similar trend in life span assay when compared to ptub::*E(z)*^*WT*^ (**FIGURE4C’, Median Survival: *E(z)***^***WT***^ **= 58, *E(z)***^***A691G***^ ***=* 51**, p<0.0001). p-value were determined by unpaired t-tests.

Given that *EZH1* is involved in the regulation of developmental target genes and that *E(z)* fly mutants display early embryonic lethality with very distinct chromatin abnormalities (O’Dor et al. 2006), we were interested in neurological and developmental phenotypes. Also, it has been observed that flies with underlying bang sensitivity have reduced life spans (Reynolds 2018). To study these in flies, we used the commonly assessed lifespan assay. When we expressed the ptub::*E(z)*^*A691G*^ transgene in the fly null background, [*ptub::E(z)*^*A691G*^*/+*; *E(z)*^*731*^*/E(z)*^*63*^] flies showed reduced longevity when compared to wild-type flies ptub::*E(z)*^*WT*^ [*ptub::E(z)*^*WT*^*/+*; *E(z)*^*731*^*/E(z)*^*63*^] (**FIGURE 4C**, p<0.0001). However, when the same transgenes were tested in the fly wild-type background, the ptub::*E(z)*^*A691G*^ [*ptub::E(z)*^*A691G*^*/+*] flies showed a similar reduction in life span than ptub::*E(z)*^*WT*^ [*ptub::E(z)*^*WT*^*/+*] (**FIGURE 4C’**, p<0.0001).

The increased bang sensitivity and reduced lifespan of the *E(z)*^*A691G*^ flies compared to the wild-type flies suggests a partial loss of function.

## DISCUSSION

Here we report a *de novo EZH1* missense variant identified in an individual with severe global developmental delay, proximal muscle weakness, acquired short stature, and dysmorphic features. While this represents an n=1 case, the evidence for pathogenicity includes 1) a *de novo* variant with damaging predictions that is absent from large population databases and lack of another compelling gene candidate, 2) the fact that the identical variant in the human paralog *EZH2* is a known disease-causing variant in patients with Weaver syndrome (Diets et al., 2018), and 3) *Drosophila* studies show the variant leads to increased H3K27 trimethylation and impairs behavior and longevity. Indeed, *de novo* Weaver syndrome missense variants result in increased H3K27 trimethylation (Cohen et al. 2015; McCabe et al. 2012)

The *Drosophila* model presented is within the *E(z)* homolog and could be considered an animal model for both this rare disease and for *EZH2*-related Weaver syndrome. While this is strong evidence for pathogenicity, additional patients and studies are required to determine which aspects of the human phenotype are due to *EZH1*.

We provide evidence that, with respect to H3K27me3, the *EZH1* variant produces a gain-of-function (GOF). In support of this, within the *EZH1* paralog *EZH2*, the analogous variant is implicated as a GOF (McCabe et al. 2012). Indeed, the trimethylation is increased in the presence of the variant in null, wild-type, and heterozygous backgrounds. The variant in the wild-type background is also bang-sensitive at 16 days, while the reference is not. However, the *in vivo* data is more complex and may suggest the *EZH1* variant is a loss of function. The variant fully rescues fly null lethality; however, the resulting flies are bang-sensitive and live a shorter lifespan than the WT rescue. This might suggest the variant is a hypomorph. It is possible that there are multiple functions of EZH1 and that the bang sensitivity and survival phenotypes are not completely coupled to the hyper-trimethylation phenotypes, which would explain the differences in function in the different assays. Alternatively, the unintended repression of gene targets could explain the loss-of-function mechanism in this scenario. The *Drosophila* model is an important *in vivo* tool for looking at variants in this complex beyond their direct effect on methylation.

Weaver syndrome is primarily known as an overgrowth syndrome, with increased height and weight, macrocephaly, and large hands. In contrast, our patient has low normal height with normal weight and head circumference, and he has small hands and feet. Retrognathia is a common feature in Weaver syndrome, while our patient had mild prognathia. Our patient is also not known to have any of the skeletal features described in Weaver syndrome. Our patient’s developmental delay and hypotonia are more severe than that is typically seen in Weaver syndrome. In addition, muscle weakness and hypopigmentation have not been described in Weaver syndrome, to our knowledge. The shared features between Weaver syndrome and our patient are relatively non-specific. These include developmental delay, hypotonia, strabismus, inverted nipples, and inguinal hernia.

We hypothesize that the differing phenotypes between Weaver syndrome and our patient may result from differences in expression patterns between *EZH2* and *EZH1. EZH2* is most abundant in proliferating tissues, particularly EBV-transformed lymphocytes, while *EZH1* is most highly expressed in the cerebellum and tibial nerve (GTEx, and (Margueron et al. 2008)). Compared to *EZH2, EZH1* is also more highly expressed in the cerebral cortex, the frontal cortex, and skeletal muscle (GTEx), consistent with our patient’s more severe neurodevelopmental phenotype and muscle weakness. The timing of expression in the developing brain also differs between the two genes. *EZH2* expression in the brain is moderately high in early the first trimester of pregnancy and then trends downward, with very low expression from late gestation through 40 years of age (BrainSpan Developmental Transcriptome). By contrast, *EZH1* expression in the brain is relatively low is the early first trimester but increases steadily throughout development (BrainSpan Developmental transcriptome). This suggests that *EZH1* may be maintaining the methylation after initial embryonic development. Thus, the *EZH1* variant identified in this patient could suppress critical genes during later stages of the embryonic development.

Undiagnosed diseases present opportunities to identify novel genetic determinants of disease. Members of the PRC2 complex have well-reported disease associations, including the A677 amino acid of the EZH2 protein. Here the equivalent amino acid of the alternative subunit and paralog, *EZH1* is identified in an undiagnosed individual with a neurodevelopmental phenotype. *Drosophila* models of the variant show hyper-trimethylation and resultant behavioral phenotypes. This model is incidentally also relevant to Weaver syndrome. Further studies will be needed to delineate the full human phenotypic spectrum for *EZH1*-related disorder.

## Materials and methods

### Ascertainment and Ethics

The patient was ascertained through participation in the Undiagnosed Diseases Network (UDN), which was approved by the National Institutes of Health Intramural Institutional Review Board. Informed consent was obtained for the proband and his first-degree relatives. The proband’s parents consented to the publication of his identifiable photos.

### Genome Sequencing and Analysis

Clinical quad genome sequencing was performed by HudsonAlpha Clinical Services Lab as part of the family’s participation in the UDN study. Genomic DNA was extracted from blood and, after quality control and fragmenting, sequenced using the Illumina HiSeqX platform to generate 150bp paired-end reads. The reads were then aligned to GRCh37/hg19 (BWA-mem v0.7.12). After quality control steps, including removal of fragments mapping to multiple locations, duplicate fragments, and fragments with low quality scores (SMAbamba v0.5.4, markdup, GATKv3.3), variant calling was performed (GATKv3.3). Variant annotation and prioritization were performed using CarpeNovo v6.0.1 and/or Codicem, a custom software analysis application. The pipeline used is expected to yield 40x coverage of 90-95% of the human reference genome.

Research-based analysis of the data by HudsonAlpha and by Brigham Genomic Medicine (PMID: 30131872) independently identified the de novo *EZH1* variant as a candidate. This variant was confirmed via dideoxy (Sanger) sequence analysis by HudsonAlpha.

### Fly Stocks

1. *E(z)*^*731*^ from Bloomington Stock Center *#*24470 *w*^*\**^; *E(z)*^*731*^ *p{1xFRT*.*G}2A/TM6C,Sb*^*1*^ *Tb*^*1*^.
2. *E(z)*^*63*^ kindly gifted by Dr. Richard Jones, Dallas, Texas: *sc z*^*1*^ *wis; E(z)*^*63*^ *e*^*11*^*/TM6B, Tb Hu*.
3. *E(z)RNAi* ordered from Bloomington Stock Center #36068 *y[1] sc[*] v[1] sev[21]; P{y[+t7*.*7] v[+t1*.*8]=TRiP*.*GL00486}attP40*
4. *p{UAS-lacZ}* gift from Hugo Bellen Lab
5. *p{Actin-GAL4}* balanced over CyO-Tb was generated in our lab.
6. *p{GawB}elav[C155]* gift from Hugo Bellen Lab
7. *p{daughterless-GAL4}* gift from Hugo Bellen Lab
8. *p{nubbin-GAL4}* gift from Hugo Bellen Lab
9. *y w; ptub::E(z)*^*WT*^*/SM6a* (generated in the lab)
10. *y w; ptub::E(z)*^*A691G*^*/SM6a* (generated in the lab)

### Fly husbandry

Stocks were reared on standard fly food (water, yeast, <u>soy flour</u>, cornmeal, agar, molasses, and propionic acid) at room temperature (∼22°C) and routinely maintained. Unless otherwise noted, all flies used in experiments were grown in a temperature and humidity-controlled incubator at 25°C and 50% humidity on a 12-hour light/dark cycle.

### Generation of transgenic flies

#### Generation of the *E(z)*^*WT*^ and *E(z)*^*A691*^ transgenic flies under UAS promoter (Supplemental Figure1)

Original E(z)-cDNA construct – pGEX-2T{E(z)cDNA e32} was kindly gifted by Dr. Richard Jones, University of Dallas (Jones and Gelbart 1990). The attB-E(z) primers were designed to clone into the *pENTR-221* vector with the help of Gateway™ BP Clonase™ II Enzyme mix (Thermo Fischer-11789100). The variant primers were prepared, and the variant was generated with a Q5 mutagenesis (NEB-E0554S) as previously explained in (Tsang et al. 2016). After confirming via sequencing analysis, *pENTR-221 E(z)cDNA*^*WT*^ and *pENTR-221-E(z)*^*A691G*^ were cloned into pGW.attB vector by using Gateway™ LR Clonase™ II Enzyme mix (Thermo Fischer-11791020). Sequence-verified wild-type and variant constructs were then microinjected in *VK00037 attP* docking site embryos to generate *pBac{UASg-E(z)*^*WT*^*}VK00037* and *pBac{UASg-E(z)*^*A691G*^*}VK00037* transgenic lines.

#### Generation of the *EZH1* transgenic flies under UAS promoter (Supplemental Figure 2)

Human cDNA “<u>IOH10021</u>” from the Ken Scott collection was used for these constructs. By designing the site-specific primers, the variant was generated with a Q5 mutagenesis protocol (NEB-E0554S) and after confirming via Sanger sequencing analysis, both the *pENTR-221-EZH1*^*ref*^ and *pENTR-221-EZH1*^*A678G*^ constructs were cloned into *pGW-{UASg}vector* by using LR Clonase™ II (Thermo Fischer-11791020). Sequence-verified *pattB{UAS-EZH1*^*ref*^*} & pattB{UAS-EZH1*^*A678G*^*}* constructs were microinjected into *VK00033* docking site embryos to generate *pBac{UAS-EZH1*^*ref*^*}VK00033 & pBac{UAS-EZH1*^*A678G*^*}VK00033*.

#### Generation of the *E(z)*^*A691*^ transgenic flies under the constitutively active tubulin promoter (Supplemental Figure 3)

*pwmc{ptub:EGFP::E(z)}* construct and flies were gifted by Dr. Leonie Ringrose, Professor, Humboldt-Universität zu Berlin, IRI Life Sciences, Berlin, Germany. The wildtype construct underwent mutagenesis. The variant was generated with a Q5 mutagenesis protocol (NEB-E0554S). Both wild-type and variant constructs underwent restriction digestion with NotI (NEB-R3189S) and XbaI (NEB-R0145S) and then ligated into the pattB-Basler vector with the T4 DNA ligase (NEB M0202S). Sequenced verified *pattB{ptub:EGFP::E(z)*^*WT*^*}*, and *pattB{ptub:EGFP::E(z)*^*A691G*^*}* then microinjected in *ϕc31* mediated *VK00037* docking site embryos to generate *pBac{ptub:EGFP::E(z)*^*WT*^*}VK00037* and *pBac{ptub:EGFP::E(z)*^*A691G*^*}VK00037* transgenic flies.

#### Longevity assay

The ptub::*E(z)*^*WT*^ and the ptub::*E(z)*^*A691G*^ transgenic flies were crossed to *y w* to avoid any phenotypic interference because of the balancer - SM6a-CyO. The F1 flies were then raised at 25^0^C and transferred every 3-4 days. Rescued flies were also treated using the same technique. Any lethality observed was plotted in Prism software (Survival plot).

### Bang sensitivity assay

Flies to be used were isolated 1-3 days post-eclosure and were separated individually for days until assessment. On the day of the trial, flies were transferred into an empty polystyrene vial. The flies were vortexed at full speed (Fisher STD Vortex Mixer, Cat. No, 02215365) for 10 seconds, and recovery times were recorded using a digital stopwatch. Bang assay was performed on a minimum of 20 flies.

### Over-expression assay (Assessment of lethality and morphological phenotypes)

Lethality and morphological phenotyping assays were performed by crossing *GAL4* drivers as indicated in the text, using 5-10 virgin females crossed to a similar number of males. Parents were transferred into a new vial after every 3-5 days to collect multiple F1-progenies. Flies were collected after most pupae eclosed, and the total number of flies was scored based on the presence or absence of balancers. For the lethality assessment, a minimum of 70 flies were scored. Viability was calculated via evaluation of the number of observed progenies compared to the number of expected progeny based on the mendelian ratio. Animals were classified as lethal if the O/E ratio was less than 0.15, and semi-lethality is classified as an O/E ratio less than 0.8. Assessment of morphological phenotypes was only done for animals lacking balancers, and phenotypes were noted if they appeared in more than 70% of the progeny.

### Western blot assay

Six adult heads per genotype (n≥3males and 3 females) or single whole adults were used (n≤10) and transferred directly to ice-cold sample buffer (4x XT Sample Buffer Bio-Rad #1610791 + 10% Beta-mercaptoethanol + ddH_2_O). The heads/whole flies were homogenized for until crushed properly, transferred to ice, and boiled at 94^0^C for 5 minutes. The boiled homogenate was centrifuged for 10 minutes at 14,000 RPM to settle the debris. Protein (10 uL) was loaded on tris-glycine precast gels (4-20% precast TGX gels Bio-Rad# 4561094). PVDF membrane was activated by applying 100% methanol for 2-3 seconds. The gels were transformed into the 0.2um activated PDVF membrane using the trans-blot turbo transfer unit. This membrane was then blocked using a 5% blocking solution (1x TBST with 0.1% TWEEN-20 and 2.5g nonfat dry milk) for 1 hour at room temperature with rotation. It was then incubated overnight at 4°C with Tri-Methyl-Histone H3 (Lys27) (CST #9733, 1:2000) or Histone H3 Antibody (CST #9715, 1:5000) primary antibodies in the blocking solution. The membrane was washed at least three times with 0.1%TBST before the secondary antibody incubation - Goat anti-Rabbit IgG (H+L) secondary Antibody, HRP (Thermo fisher #31460, 1:5000 in the blocking solution). SuperSignal™ West Pico PLUS (34580, Thermo) to develop the membrane. Images of the developed blot were taken in the Chemiblot imager and were then analyzed using the ImageJ software.

### ver-expression assay

*UAS-E(z)*^*WT*^ and *UAS-E(z)*^*A691G*^ transgenic flies were crossed with different tissue-specific and ubiquitous *GAL4* lines and grown at different temperatures (explained in tables).

All Drosophila strains not already available in the Bloomington stock center are upon request. The authors affirm that all the data necessary for testing the assertions in this manuscript are shown in the manuscript and data presented.

### Websites accessed

DenovoDB: <u>denovo-db.gs.washington.edu</u>

GETx: https://gtexportal.org/home/

## Data Availability

All data produced in the present study are available upon reasonable request to the authors.

## Acknowledgments

This paper is supported by NIH grants U54 NS 093793 (The UDN MOSC) to M.W and S.Y U01HG007690,, 3U01HG007690-08S1 (D.A.S., M.A.W, F.A.H, L.C.B). The Translational Clinical Research Center at Massachusetts General Hospital was supported by NIH grant number 1UL1TR001102. The Hill family Fund for the Diagnosis, Management of Rare and Undiagnosed Diseases and the American Institute for Neuro-Integrative Development (AIND) at Mass General, and Brigham Research Institute Director’s Transformative Award to Brigham Genomic Medicine also supported this work. We are thankful to Dr. Leonie Ringrose, Berlin, Germany, and Dr. Richard Jones, Dallas, Texas USA, for kindly proving the cDNA and the fly reagents for the study. Also, we are thankful for the resources provided by the Bloomington *Drosophila* Stock Center, and to Ken Scott collection for generous donation of the EZH1 human cDNA plasmid.

## Supplementary data

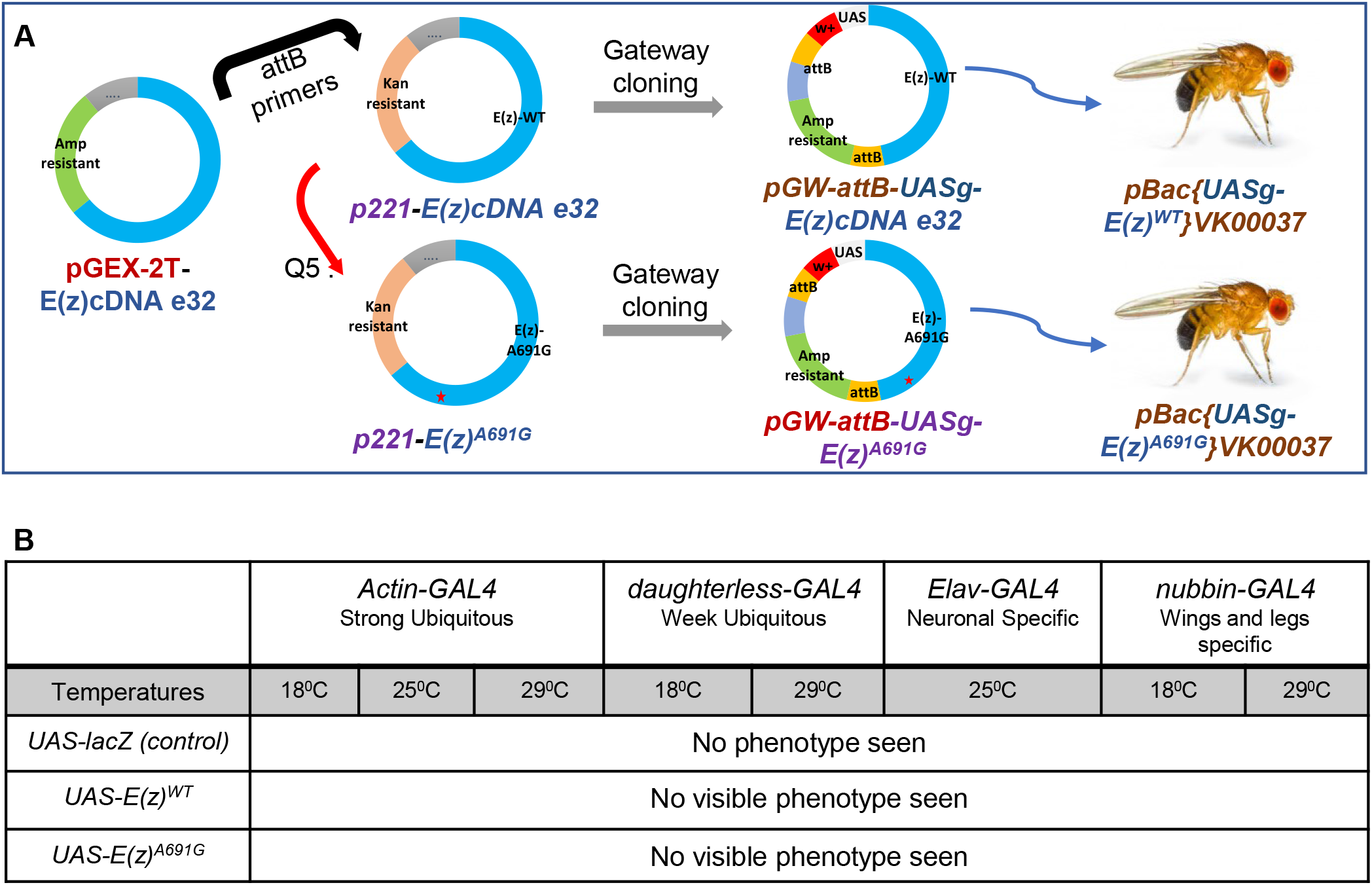

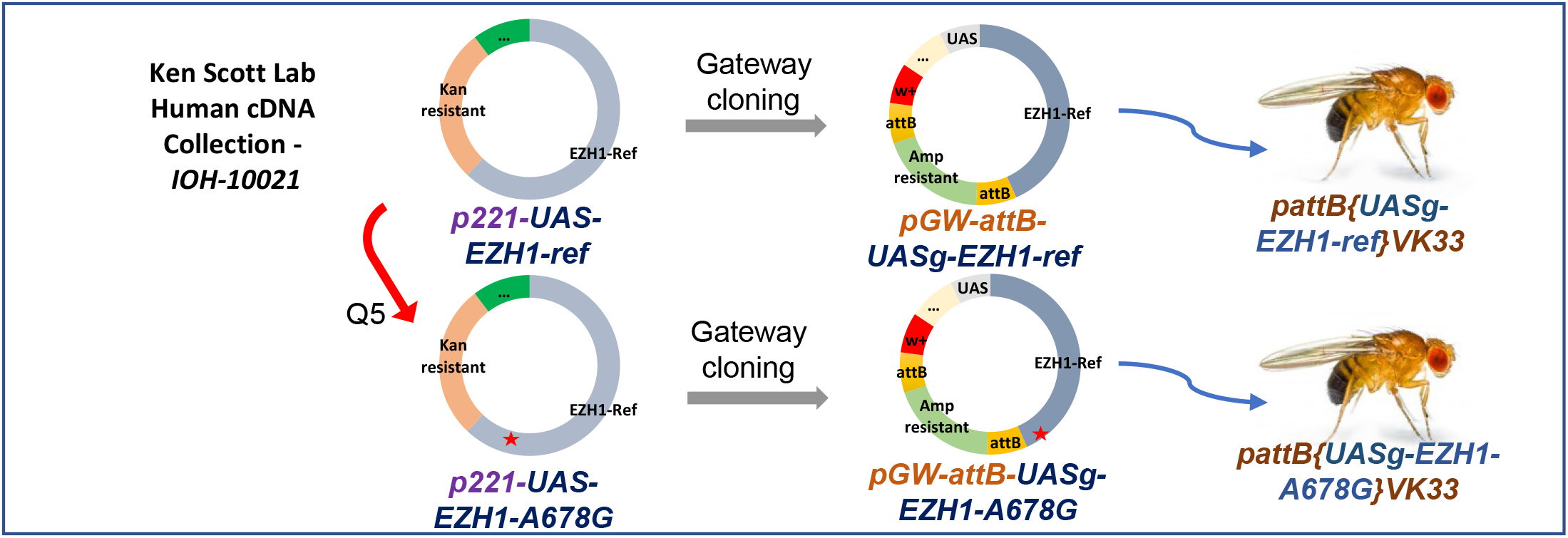

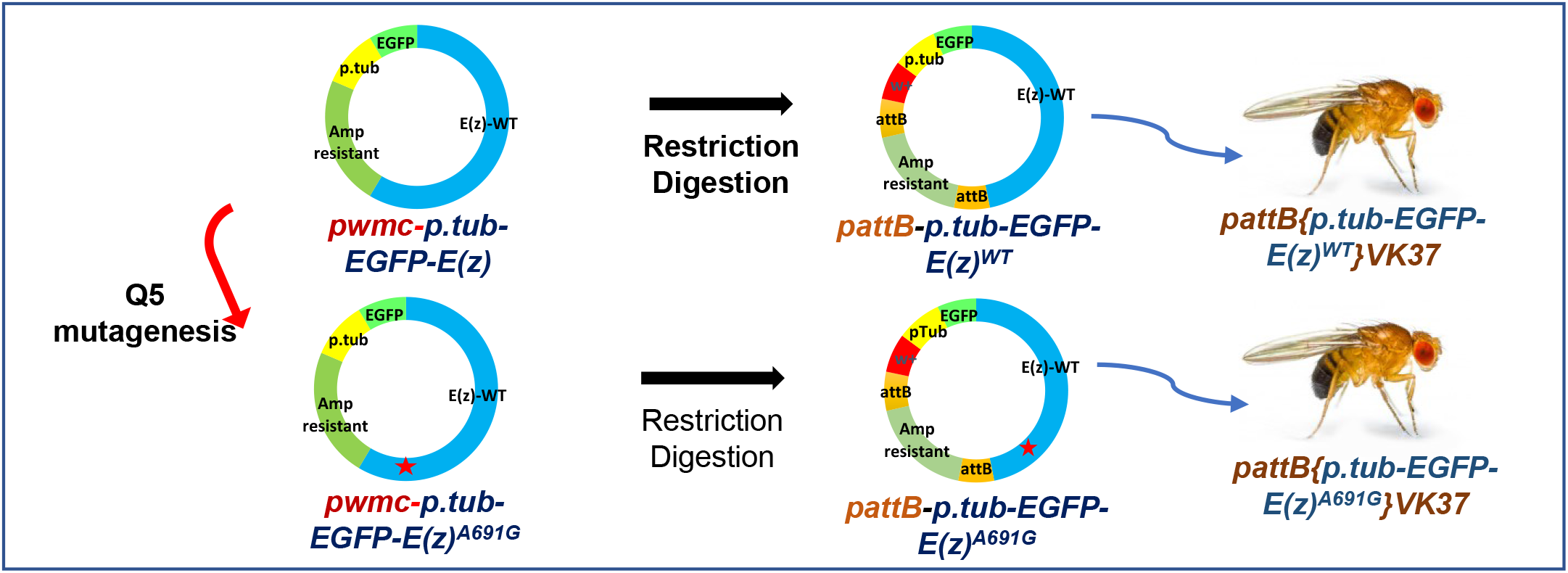

**Supplementary Figure1:**
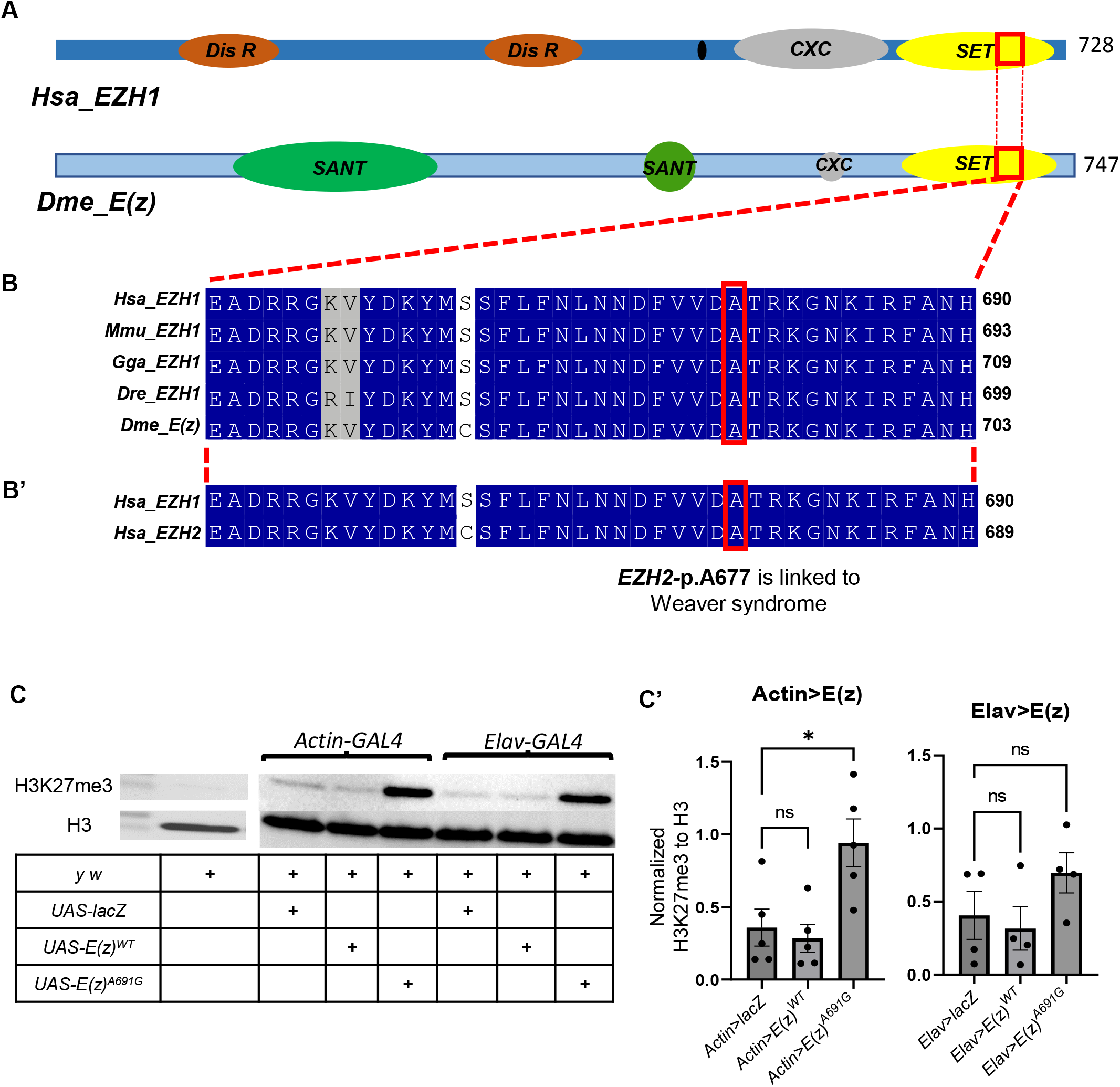
Generation of the *E(z)*^*A691*^ transgenic flies under the UAS-promotor. Original E(z)-cDNA construct – pGEX-2T{E(z)cDNA e32} was kindly gifted by Dr. Richard Jones, University of Dallas (Jones and Gelbart 1990). The attB-E(z) primers were designed to clone into the pENTR-221 vector with the help of Gateway™ BP Clonase™ II Enzyme mix (Thermo Fischer-11789100). The variant primers were prepared, and the variant was generated with a Q5 mutagenesis (NEB-E0554S) as previously explained in (Tsang et al. 2016). After confirming via sequencing analysis, pENTR-221 E(z)cDNAWT and pENTR-221-E(z)A691G were cloned into pGW.attB vector by using Gateway™ LR Clonase™ II Enzyme mix (Thermo Fischer-11791020). Sequence-verified wild-type and variant constructs were then microinjected in VK00037 attP docking site embryoes to generate pBac{UASg-E(z)WT}VK00037 and pBac{UASg-E(z)A691G}VK00037 transgenic lines. **Table B**: **Overexpression assay** performed with different GAL4 lines at different temperatures.

**Supplementary Figure2:**
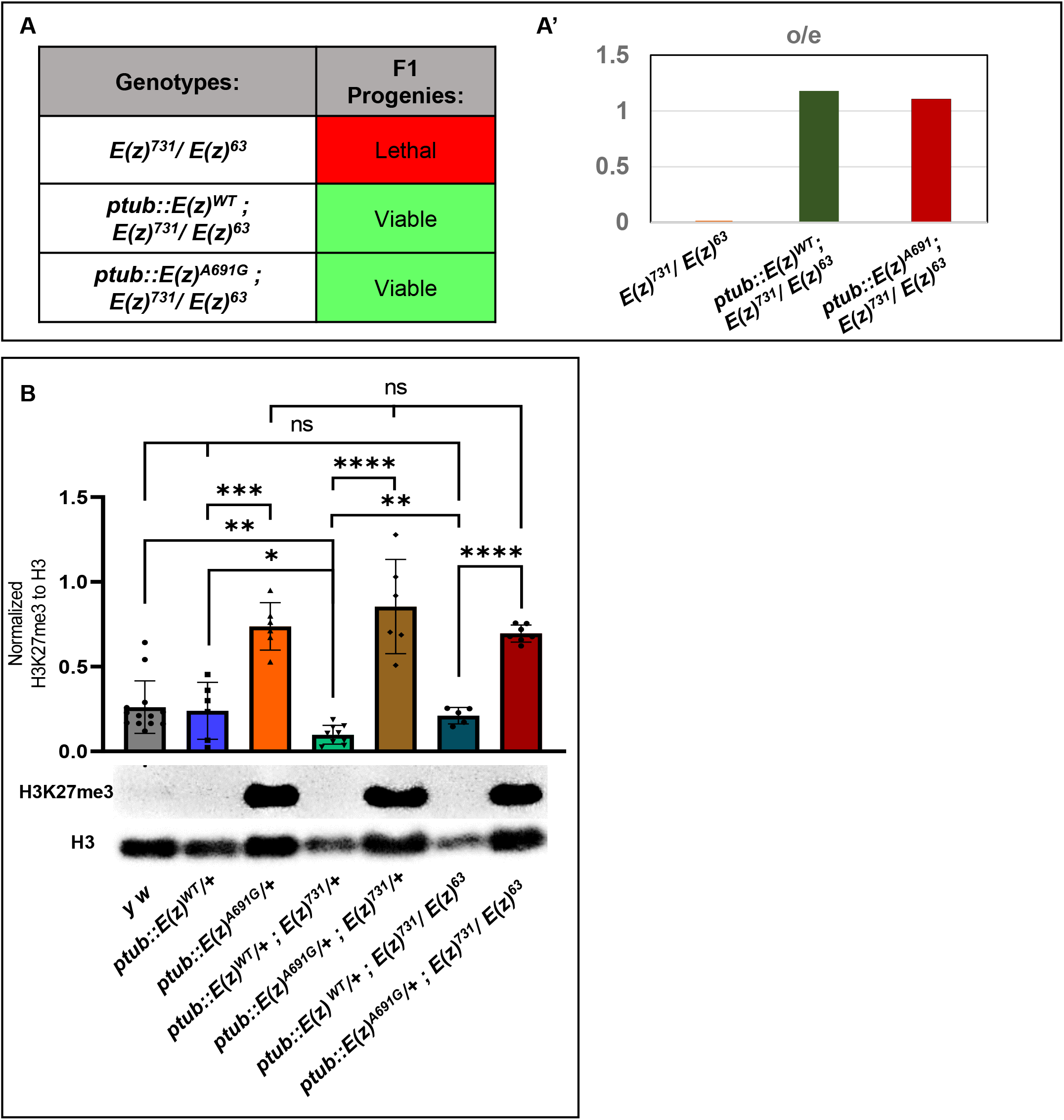
Generation of the human *EZH1-cDNA* transgenic flies under UAS promotor. Human cDNA “IOH10021” from the Ken Scott collection was used for these constructs. By designing the site-specific primers, the variant was generated with a Q5 mutagenesis protocol (NEB-E0554S) and after confirming via Sanger sequencing analysis, both the pENTR-221-EZH1ref and pENTR-221-EZH1A678G constructs were cloned into pGW-{UASg}vector by using LR Clonase™ II (Thermo Fischer-11791020). Sequence-verified pattB{UAS-EZH1ref} & pattB{UAS-EZH1A678G} constructs were microinjected into VK00033 docking site embryos to generate pBac{UAS-EZH1ref}VK00033 & pBac{UAS-EZH1A678G}VK00033.

**Supplementary Figure3:**
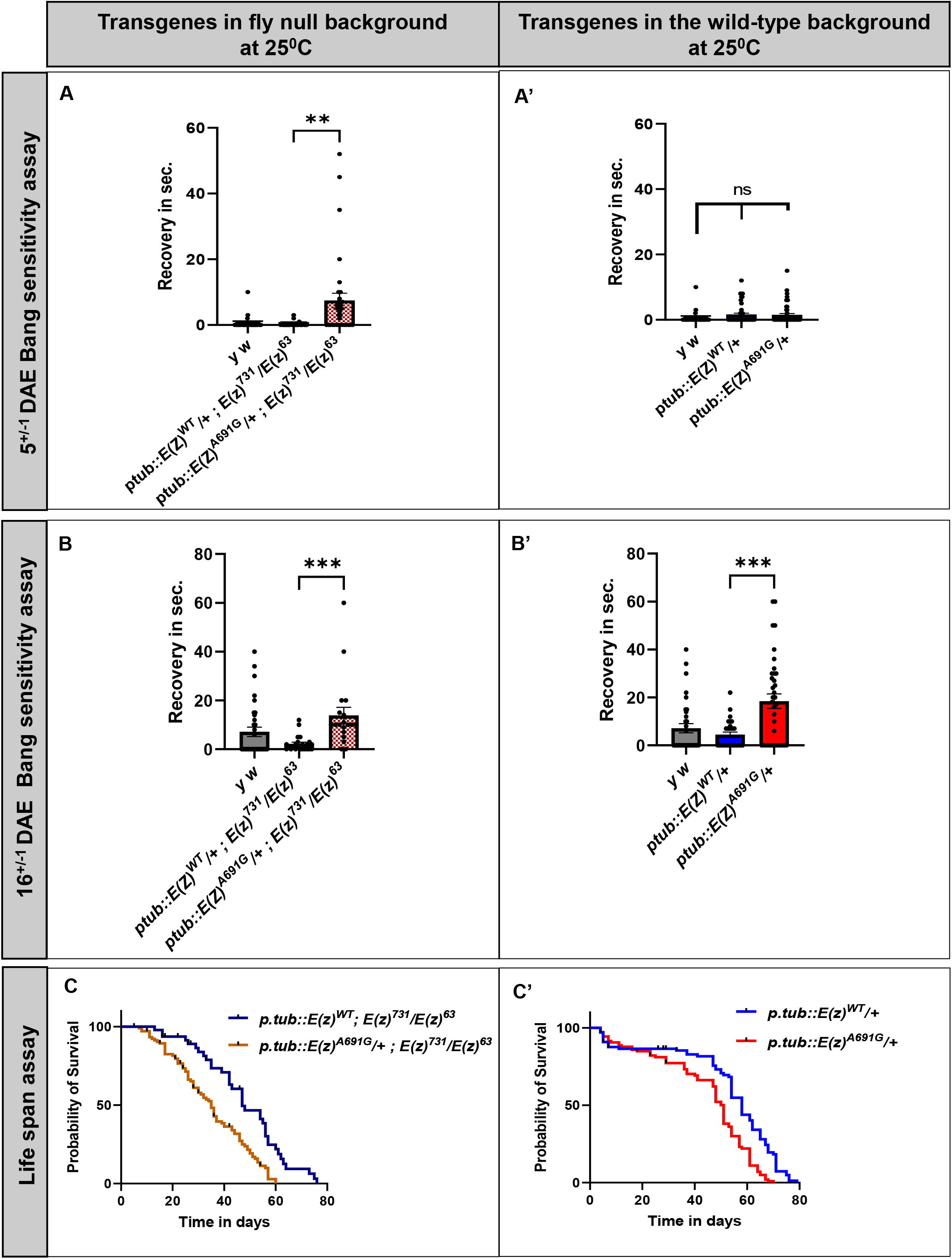
Generation of the *E(z)*^*A691*^ transgenic flies under the constitutively active tubulin promotor. *pwmc{ptub:EGFP::E(z)}* construct and flies were gifted by Dr. Leonie Ringrose, Professor, Humboldt-Universität zu Berlin, IRI Life Sciences, Berlin, Germany. The wildtype construct underwent mutagenesis. The variant was generated with a Q5 mutagenesis protocol (NEB-E0554S). Both wild-type and variant constructs underwent restriction digestion with NotI (NEB-R3189S) and XbaI (NEB-R0145S) and then ligated into the pattB-Basler vector with the T4 DNA ligase (NEB M0202S). Sequenced varified *pattB{ptub:EGFP::E(z)*^*WT*^*}*, and *pattB{ptub:EGFP::E(z)*^*A691G*^*}* then microinjected in ϕc31 mediated *VK00037* docking site embryos to generate *pBac{ptub:EGFP::E(z)*^*WT*^*}VK00037* and *pBac{ptub:EGFP::E(z)*^*A691G*^*}VK00037* transgenic flies.

