## Supplementary figure 1 for "A *de novo* missense variant in *EZH1* associated with developmental delay exhibits functional deficits in *Drosophila melanogaster*"

### Supplementary data

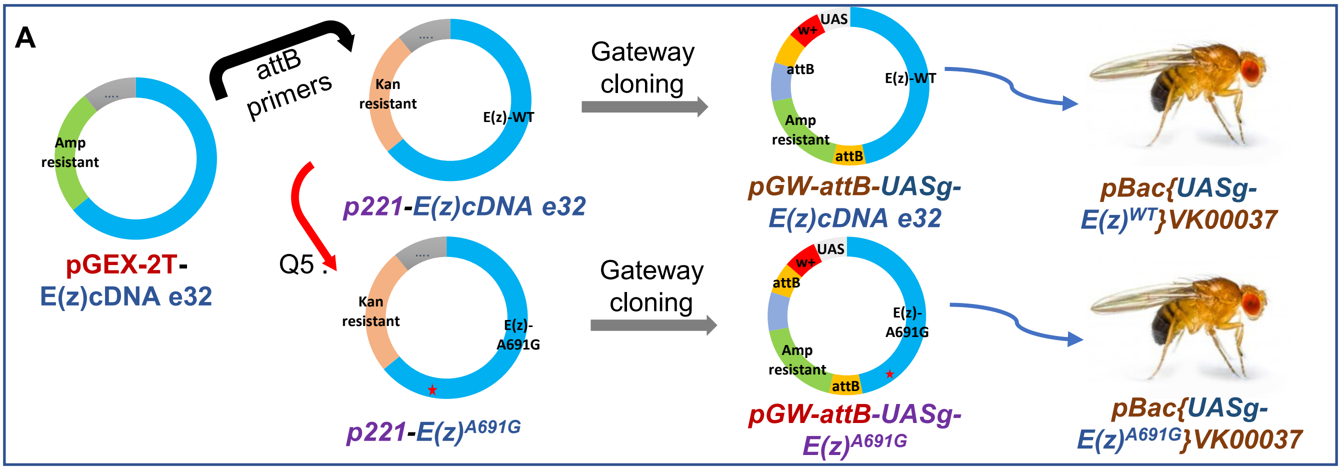


**B**

| | *Actin-GAL4* Strong Ubiquitous | | | *daughterless-GAL4* Week Ubiquitous | | *Elav-GAL4* Neuronal Specific | *nubbin-GAL4* Wings and legs specific | |
|---|---|---|---|---|---|---|---|---|
| Temperatures | 18$^0$C | 25$^0$C | 29$^0$C | 18$^0$C | 29$^0$C | 25$^0$C | 18$^0$C | 29$^0$C |
| *UAS-lacZ (control)* | No phenotype seen | | | | | | | |
| *UAS-E(z)$^{WT}$* | No visible phenotype seen | | | | | | | |
| *UAS-E(z)$^{A691G}$* | No visible phenotype seen | | | | | | | |
