## Supplementary figures and images for "A *de novo* missense variant in *EZH1* associated with developmental delay exhibits functional deficits in *Drosophila melanogaster*"

### Supplementary figure 2

#### Supplementary data

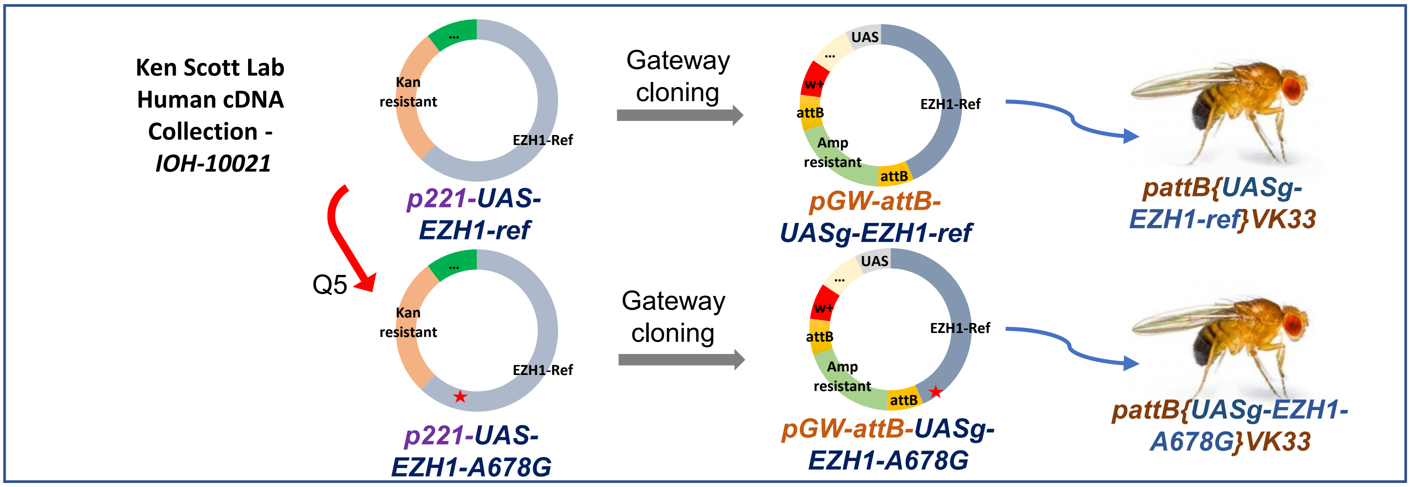

### Supplementary figure 3

#### Supplementary data

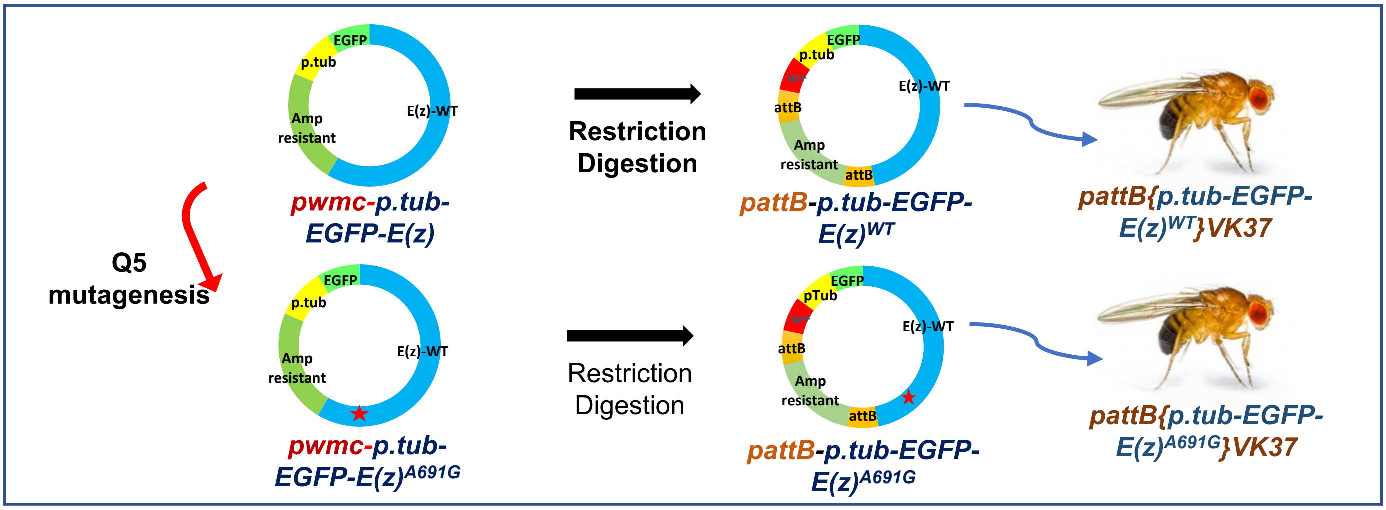
